# Metabolomic profiles of chronic distress predict future cardiovascular disease risk

**DOI:** 10.1101/2022.02.26.22271549

**Authors:** Raji Balasubramanian, Katherine H. Shutta, Marta Guasch-Ferre, Tianyi Huang, Shaili C. Jha, Yiwen Zhu, Aladdin H. Shadyab, JoAnn E. Manson, Frank Hu, Kathryn M. Rexrode, Clary B. Clish, Susan E. Hankinson, Laura D. Kubzansky

## Abstract

**Background:** Chronic psychological distress has been associated with increased risk of cardiovascular disease (CVD). However, mechanistic evidence explaining the observed associations remains limited and, with data are particularly sparse among women. This study examined if a metabolite profile linked with distress would be associated with increased risk of CVD.

**Methods:** A plasma metabolite-based distress score (MDS) of twenty metabolites was derived in a cross-sectional, 1:1 matched case-control dataset (n=558 women) in the Nurses’ Health Study (NHS). We then calculated this score in two other cohorts, the Women’s Health Initiative Observational Cohort (WHI-OS) and the Prevención con Dieta Mediterránea (PREDIMED) trial, and tested association with risk of developing adjudicated measures of CVD in each cohort. We considered incident coronary heart disease (CHD) in the WHI-OS dataset which included 944 postmenopausal women (472 CHD cases; mean time to event of 5.8 years), and incident CVD (including stroke, myocardial infarction, CVD death) in the PREDIMED dataset which included 980 men and women (224 CVD cases, mean time to event of 3.1 years).

**Results:** In the WHI-OS, a 1-SD increase in the plasma MDS was associated with a 14% increased risk of incident CHD (odds ratio [OR]=1.14, 95% CI: 1.03 – 1.26), adjusting for known CVD risk factors excluding total and HDL cholesterol. This association was attenuated after including total and HDL cholesterol (OR=1.09; 95% CI: 0.98 – 1.21). Of the component metabolites in the MDS, tryptophan and threonine were inversely associated with incident CHD risk. In PREDIMED, each one SD increase in the MDS was associated with a 17% increased incident CVD risk (OR=1.17, 95% CI: 1.00 – 1.38), after adjusting for risk factors including total and HDL cholesterol. Similar associations were observed in men and women. Four individual metabolites in the MDS were associated with incident CVD risk in fully adjusted models in PREDIMED. Biliverdin and C36:5 PC plasmalogen had inverse associations, whereas C16:0 ceramide and C18:0 LPE each had positive associations with CVD risk.

**Conclusions:** Our study sheds light on the key molecular alterations that characterize chronic distress and are predictive of subsequent CVD risk in men and women. These findings provide additional evidence for the role of distress in CVD development.

## Introduction

Chronic distress and related psychological conditions including depression and anxiety represent a significant public health burden, and prevalence is substantially higher in women. Moreover, chronic distress and cardiovascular disease (CVD) are established co-morbid conditions^1^. Chronic distress has been strongly linked to metabolic conditions such as diabetes and metabolic syndrome that increase risk of CVD ^2,3^, the metabolic underpinnings of this potential causal association are unclear. Substantial literature suggests depression and other forms of psychological distress are associated with substantially higher risk of CVD^4 5^. Unhealthy behaviors (e.g., unhealthy diet, sedentary behavior) as well as various biological pathways (e.g., increased inflammation, dysregulation in hypothalamic-pituitary-adrenal axis function, dyslipidemia) are hypothesized pathways underlying the association ^2,3^, but a detailed set of molecular mechanisms that could explain observed associations has not been fully delineated, and stronger mechanistic evidence is needed ^6^.

Recent work has identified numerous small-molecule metabolites whose levels differ across individuals with higher versus lower psychological distress levels ^7,8^. Identifying alterations in circulating small molecule metabolites that commonly occur not only with chronic distress but also with CVD can help shed light on the mechanisms underpinning the link between these conditions. Consistently altered metabolite levels in relation to psychological distress might serve as an important pathway linking distress to increased CVD risk. Thus, drawing on prior work identifying relevant metabolites related to distress ^7,8^, we developed a small-molecule metabolite profile of chronic distress measured in plasma and evaluated its association with incident CVD risk in multiple independent cohorts.

A number of studies have identified specific lipids and amino acids linked with depression or depressive symptoms. In our prior work, we validated many of these associations with a broader measure of distress (characterized by high levels of both anxiety and depression) and further conducted an agnostic investigation into the metabolomic fingerprint of chronic distress in studies nested within two large scale cohorts of women, the Nurses’ Health Study (NHS) and the Women’s Health Initiative (WHI) Hormone Therapy Trials ^7,8^. Several other studies have also identified metabolomic alterations associated with incident CVD risk ^9-11^. However, to our knowledge, no studies have directly examined whether any common metabolite alterations associated with chronic distress are predictive of future CVD risk.

To examine this question, we conducted a study in three stages. In Stage 1, we developed a plasma metabolomic score of chronic distress using the same data [we used for a prior distress metabolomics investigation we conducted in the Nurses’ Health Study (NHS) ^8^. While chronic distress can result from several factors including cumulative stress from daily stressors and exposure to traumatic events, and takes various forms, including feelings of chronic anger and hostility, depression, anxiety, and post-traumatic stress disorder (PTSD), for the current study we defined chronic distress according to levels of anxiety or depression, given their high prevalence in the population^12^. This score represents a cumulative summary of chronic distress as captured by an individual’s metabolomic profile. Next, we calculated this distress metabolomic score in two independent cohorts and evaluated associations of the score and its individual metabolite components with risk of incident CVD related outcomes. In Stage 2, using prospective data, we examined associations with incident coronary heart disease (CHD) risk in the Women’s Health Initiative Observational Study (WHI-OS). For added consistency, using data from WHI-OS we evaluated cross-sectional associations between the distress metabolomics score and the Framingham Risk Score ^13^. In Stage 3, we sought to replicate the findings in the Spanish Prevención con Dieta Mediterránea (PREDIMED) trial, considering incident CVD risk^14^. In both Stages 2 and 3 we additionally adjusted for key potential confounders of the association between a distress metabolite score and CVD-related outcomes, identified based on prior literature, such as medication use and lifestyle factors.

## Methods and Materials

### Study Populations

This study included participants from three different cohorts of older adults: the NHS (all women), WHI–OS (all women), and PREDIMED (women and men) (Table 1). All participants in each cohort provided written informed consent and our study was approved by the Institutional Review Board of UMass-Amherst.

**Table 1:** Basic characteristics of the NHS, WHI-OS, PREDIMED, assessed at or near time of blood collection. NA: not applicable.

|  | CHD/CVD |  |  |  | Distress |  |
| --- | --- | --- | --- | --- | --- | --- |
|  | WHI-OS |  | PREDIMED |  | NHS |  |
|  | Recruitment: 1993-1998<br>Blood collection: at study entry |  | Recruitment: 1989<br>Blood collection: 2013-2014 |  | Recruitment: 1976<br>Blood collection: 2000-2002 |  |
|  | CHD case | CHD-control | CVD case | Sub-cohort | Not Distressed | Distressed<sup>1</sup> |
| N | 472 | 472 | 224 | 756 | 279 | 279 |
| Age in years, mean(sd) | 67.5 (6.9) | 67.2 (6.8) | 69.3 (6.5) | 67.1 (5.9) | 64.3 (6.6) | 64.3 (6.6) |
| Non-white, % | 26.3 | 26.3 | 0 | 0 | 2.5 | 0.7 |
| Female, % | 100 | 100 | 39.3 | 58.3 | 100 | 100 |
| **Measures of depression** |  |  |  |  |  |  |
| CES-D-6, mean(sd)<br>*N missing* | 2.45 (2.66)<br>5 | 2.18 (2.31)<br>9 | NA | NA | NA | NA |
| CES-D-10, mean(sd) | NA | NA | NA | NA | 3.7 (2.5) | 15.6 (4.3) |
| Antidepressant use, % | 6.14 | 6.36 | NA | NA | 1.4 | 47.7 |
| History of depression, %<br>*N missing* | 13.5<br>6 | 8.8<br>6 | NA | NA | 0 | 55.2 |
| **Comorbidities** |  |  |  |  |  |  |
| Type 2 diabetes, % | 16.5 | 5.3 | 63.8 | 46.4 | 2.9 | 13.3 |
| Hypertension, %<br>*N missing* | 52.3<br>2 | 31.7<br>2 | 82.1 | 83.9 | 38.7 | 53.8 |
| **Medications** |  |  |  |  |  |  |
| Aspirin use, % | 24.6 | 20.8 | NA | NA | NA | NA |
| Statin use, % | 8.9 | 8.3 | NA | NA | 17.2 | 27.6 |
| Other lipid lowering drug,% | 1.1 | 0.4 | NA | NA | 1.8 | 5.4 |
| Hormone therapy, % | 2.5 | 2.5 | NA | NA | 62.2 | 62.3 |
| **Cardiovascular risk factors** |  |  |  |  |  |  |
| Systolic blood pressure,<br>mmHg<br>*N missing* | 135.4 (19.4) | 129.7 (18.3) | 154.7 (21.2) | 147.5 (18.7)<br>1 | NA | NA |

<sup>1</sup> Current and/or persistent depression and/or anxiety (detailed in Methods)

|  | CHD/CVD |  |  |  | Distress |  |
| --- | --- | --- | --- | --- | --- | --- |
|  | WHI-OS |  | PREDIMED |  | NHS |  |
|  | Recruitment: 1993-1998<br>Blood collection: at study entry |  | Recruitment: 1989<br>Blood collection: 2013-2014 |  | Recruitment: 1976<br>Blood collection: 2000-2002 |  |
|  | CHD case | CHD-control | CVD case | Sub-cohort | Not Distressed | Distressed<sup>1</sup> |
| Total cholesterol, mg/dL<br>*N missing* | 230.7 (47.3) | 232.7 (47.2) | 208.4 (34.0)<br>2 | 207.4 (35.8)<br>19 | NA | NA |
| HDL cholesterol, mg/dL<br>*N missing* | 50.9 (15.8) | 56.9 (17.1) | 49.4 (11.1)<br>6 | 51.2 (11.5)<br>25 | NA | NA |
| **Biobehavioral factors** |  |  |  |  |  |  |
| Body mass index, kg/m<sup>2</sup><br>*N missing* | 28.5 (6.4)<br>1 | 27.2 (5.8) | 29.6 (3.8) | 29.8 (3.6) | 25.7 (4.5) | 28.5 (7.6) |
| Current smokers, % | 10.2 | 5.9 | 20.5 | 12.3 | 4.7 | 7.9 |
| Physical activity in MET-hrs/week, mean(sd) | 11.2 (12.1) | 14.0 (14.3) | NA | NA | 25.9 (28.5) | 16.9 (22.1) |
| Diet quality score<sup>2</sup><br>mean(sd)<br>*N missing* | 67.9 (11.1)<br>2 | 69.0 (11.4) | NA | NA | 57.5 (13.1) | 53.6 (12.5) |
| Caffeine intake, mg/day<br>mean(sd)<br>*N missing* | 150.0 (133)<br>2 | 151.1 (128.3) | NA | NA | 124.1 (115.1) | 144.9 (137.9) |
| Alcohol intake, g/day<br>mean(sd)<br>*N missing* | 4.7 (9.6)<br>2 | 5.7 (13.0)<br>1 | NA | NA | 7.3 (12.0) | 5.4 (9.9) |

<sup>2</sup> Measured by HEI-2005 in WHI and AHEI-2010 in NHSII and NHS.

The NHS consists of 121,700 US registered female nurses aged 30-55 years at study enrollment in 1976^15^. The NHS includes two blood collections. Women who provided blood samples at the first blood collection (1989–1990) were invited to give a second blood sample (2000-2002) as described in detail elsewhere^16^. Participants eligible for the current study provided blood samples in the second collection, depression-related data on questionnaires from 1992 through 2004, anxiety-related data on questionnaires in 1988 and in 2004, and were free of cancer or cardiovascular disease at the time of the second blood collection (N=13,194). Among these participants, we selected a subset of 558 women for a 1:1 matched distress-related case-control study for whom we obtained metabolomic assays. Distressed cases were matched to controls on age, race/ethnicity, menopausal status, fasting status, and date/time of day of blood draw.

The WHI consists of 161,808 postmenopausal women aged 50-79 recruited at 40 clinical centers across the US from 1993 to 1998. A subset of these women participated in the Observational Study (WHI-OS, N=93,676)^17^. The current study draws on data from an ancillary 1:1 matched CHD case-control study published in 2018 that included 944 participants selected from the WHI-OS^9^. Equal numbers of incident CHD cases and healthy controls were frequency matched on 5-year age, race/ethnicity, hysterectomy status, and 2-year enrollment groups. Women provided a fasting blood sample and completed questionnaires assessing demographic characteristics, medical, and psychosocial characteristics including measures of distress at entry into WHI in the timeframe 1993-1998. We note that the data on participants in the WHI Hormone Therapy Trials were excluded in this study as they were used in our prior agnostic investigation of distress and metabolomics ^7,8^.

The Prevención con Dieta Mediterránea (PREDIMED) trial^14^ randomized 7447 men and women at high risk of developing CVD to one of 3 interventions: a Mediterranean dietary intervention supplemented with virgin olive oil, a Mediterranean dietary intervention supplemented with nuts, or receiving advice about a low-fat diet. The primary endpoint was a composite measure of myocardial infarction (MI), stroke and CVD death. A metabolomics sub-study was conducted, using a case-cohort study design^18^ nested within PREDIMED - a random selection of eligible participants with baseline EDTA plasma samples were included (approximately 10% of the cohort) along with all remaining incident CVD cases with available samples. This PREDIMED metabolomics sub-study had 980 participants, including 224 incident CVD cases ^19 20,21^.

### Outcome and covariate assessment

#### Stage 1 Developing a metabolite-based distress score

##### Primary endpoint

Chronic distress, the outcome of interest, was derived using data from the NHS. Cases were identified according to presence of severe distress. This was characterized by repeated assessments of depression status (symptoms, reports of physician diagnosis, or antidepressant use) and anxiety (symptoms) measured prior to and around the time of blood draw. Controls consisted of women who reported neither depression nor anxiety (past or current according to the assessments described above). Details on the definition of chronic distress are provided in the Supplement and in Shutta, KH et al. (2021) ^8^.

##### Matching factors

A minimal set of important potential confounders were used as matching factors in the study design as described in Shutta, KH et al. (2021) ^8^. These factors included age, race/ethnicity (white/non-white), fasting status (yes/no), menopausal status (yes/no), and date and time of blood draw. Age and race/ethnicity were self-reported at study entry and other matching factors assessed by questionnaire at the time of blood draw.

#### Stage 2. Evaluating the MDS in relation to CHD

##### Primary endpoint

Using data from WHI-OS, the primary endpoint is incident CHD, defined as myocardial infarction or death attributable to CHD^9,22^. CHD outcomes were physician adjudicated based on review of elements of the medical history, electrocardiogram readings, and results of cardiac enzyme/troponin determinations ^22^.

##### Matching factors included in study design

included self-reported data at study entry of age (continuous), race/ethnicity (American Indian, Asian/Pacific Islander/Black, Hispanic, White, unknown), hysterectomy status (yes/no), and time period for enrollment into the WHI-OS ^8^.

##### CHD risk factors

These variables were included as covariates in the models for the primary outcome (incident CHD). These variables, assessed by study staff at the baseline clinic visit, included: systolic blood pressure (SBP), waist-to-hip ratio (WHR, continuous) and body mass index (BMI, kg/m^2^). Measurements of C reactive protein (CRP, continuous), total cholesterol (continuous) and HDL cholesterol (continuous) were obtained from fasting blood samples obtained at study entry. Details of these measures can be found in the supplement. We also calculated the Framingham Risk Score, following the published sex-specific algorithm for 10-year CVD risk.^13^ The components of the score for both men and women include age, diabetes, current smoking status, treated or untreated SBP, total cholesterol and HDL cholesterol.

##### Potential confounders/other covariates

Assessed at study entry, these covariates include: medication use [hypertension treatment (yes/no), diabetes treatment (yes/no), aspirin use (yes/no), statin use (yes/no)], co-morbidities [diabetes (yes/no), hypertension (yes/no)], smoking status (never/past/current), diet quality (Healthy Eating Index (HEI-2005) score^23^, continuous), alcohol (drinks/day) and physical activity level (MET-hours/week). Details are provided in the Supplement.

#### Stage 3. Replicating the MDS-CVD association in an independent cohort

##### Primary endpoint

Using data from the PREDIMED study, the primary endpoint was CVD, defined as a composite of myocardial infarction, stroke, and death from cardiovascular causes^14^. Details of the assessment of the endpoint have been previously described^14^. Briefly, incident CVD cases were determined using repeated contact with participants, contact with family physicians, annual review of medical records, and the National Death Index.

##### CHD risk factors

As in the WHI-OS, we included these variables described below as covariates in the models for the primary outcome (incident CVD). In addition, these factors were used to calculate the Framingham Risk Score stratified by sex for 10-year cardiovascular disease risk, considered as a secondary outcome in subsequent analyses. ^13^ Relevant covariates included randomized trial arm ^14^, age (continuous), sex, smoking status (current smoker vs other) and co-morbidities including elevated cholesterol (yes/no), hypertension (yes/no), diabetes (yes/no) collected from participants by the study nurse at the baseline visit. At the same visit, blood pressure (SBP, continuous) was measured by the study nurse and blood samples were collected from which total cholesterol (continuous) and HDL cholesterol (continuous) measurements were made.

### Metabolomics assay

For all cohorts plasma metabolomics profiling was conducted at the Broad Institute using the liquid chromatography-tandem mass spectrometry (LC-MS/ MS) method. Complementary LC-MS methods were used in all three metabolomic studies and quality control procedures that are standard for the Broad metabolomics platform were followed. For each method, pooled plasma reference samples were included every 20 samples and results were standardized using the ratio of the value of the sample to the value of the nearest pooled reference multiplied by the median of all reference values for the metabolite. For each method, metabolite identities were confirmed using authentic reference standards or reference samples. Blinded split samples were also included and used to calculate the CV for each metabolite (Table 2). Details regarding the metabolomics platform are in the Supplement and in previous publications ^8,9,14^

**Table 2:** Metabolites included in the 20-metabolite-based distress score (20-MDS) and 17-metabolite-based distress score (17-MDS). Regression coefficients were estimated in the NHS in conditional logistic regression models adjusting for matching factors.

| *Metabolite* | *HMDB* | *20-MDS*<br>*Coefficient* | *17-MDS Coefficient* | *CV in NHS* |
| --- | --- | --- | --- | --- |
| *serotonin* | HMDB0000259 | -0.59 | - | 12.07 |
| *N2,N2-dimethylguanosine* | HMDB0004824 | 0.57 | - | 15.92 |
| *threonine* | HMDB0000167 | -0.44 | -0.25 | 5.86 |
| *hippurate* | HMDB0000714 | -0.43 | -0.42 | 9.01 |
| *biliverdin* | HMDB0001008 | -0.42 | -0.47 | 21.28 |
| *C34:3 PC* | HMDB0008006 | -0.40 | -0.47 | 11.77 |
| *glutamine* | HMDB0000641 | 0.37 | 0.34 | 7.25 |
| *cotinine* | HMDB0001046 | 0.37 | 0.20 | 9.88 |
| *creatinine* | HMDB0000562 | -0.36 | -0.22 | 4.08 |
| *arachidonate* | HMDB0001043 | -0.33 | - | 9.89 |
| *3-methylxanthine* | HMDB0001886 | 0.33 | - | 9.29 |
| *C38:3 PC* | HMDB0008047 | 0.31 | 0.10 | 10.7 |
| *tryptophan* | HMDB0000929 | -0.25 | -0.36 | 6.88 |
| *GABA* | HMDB0000112 | -0.15 | -0.01 | 6.36 |
| *C16:0 Ceramide (d18:1)* | HMDB0004949 | 0.13 | 0.29 | 12.93 |
| *N4-acetylcytidine* | HMDB0005923 | -0.12 | 0.00 | 10.09 |
| *C36:5 PC plasmalogen-B* | HMDB0011220 | -0.11 | -0.18 | 8.51 |
| *C18:0 LPE* | HMDB0011130 | 0.11 | 0.18 | 11.79 |
| *pseudouridine* | HMDB0000767 | 0.05 | 0.18 | 7.83 |
| *thiamine* | HMDB0000235 | 0.05 | 0.09 | 9.94 |
| *DMGV* | HMDB0240212 | - | 0.36 | 18.3 |

### Statistical Analysis

#### Overview of Analytic Strategy

The analysis proceeded in three main stages. First, in the NHS, we selected metabolites for inclusion into the metabolite-based distress score (MDS) and derived their coefficients based on the association with chronic distress (Figure 1). Second, the MDS was calculated for each participant in the WHI-OS and tested for association with incident CHD. Third, we tested if the association of the MDS with incident CVD was replicated in PREDIMED; within each of these cohorts, the association of the MDS with incident CHD (WHI-OS) and incident CVD (PREDIMED) was evaluated after adjusting for known CHD/CVD risk factors. Each of these three stages are described in detail below.

**Figure 1:**
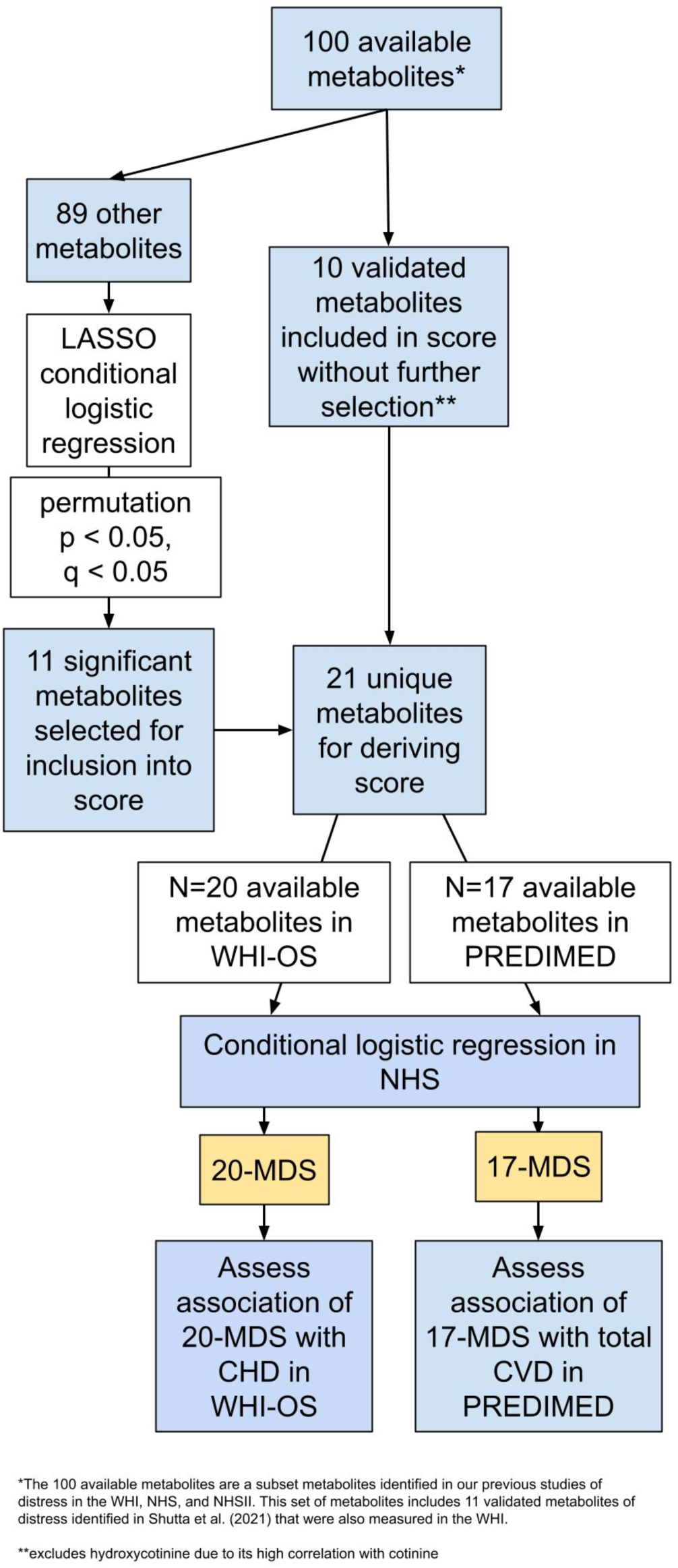
Analytical approach for deriving metabolite-based distress scores (20-MDS, 17-MDS) in the NHS and assessing association with CHD in the WHI-OS and total CVD in PREDIMED.

#### Stage 1: Deriving MDS in NHS

We cast a broad net for selecting metabolites as candidates for inclusion into the MDS by considering three categories of metabolites with evidence of association with chronic distress, based on our previous research:

(a) 8 metabolites with associations with chronic distress that met a threshold of nominal p < 0.05 in a previous meta-analysis of candidate lipids and amino acids^7^

(b) 46 metabolites we previously identified in the discovery stage of an agnostic analysis in the discovery-validation workflow presented by Shutta, KH et al. (2021)^8^

(c) 102 metabolites identified in an agnostic analysis in the NHS chronic distress case-control study (n=558, 279 chronic distress cases) which satisfied a threshold of p value < 0.05 and q value < 0.20 in conditional logistic regression models adjusting for matching factors. In summary, these overlapping metabolite sets (a)-(c) included 100 unique metabolites, which were divided into two categories for inclusion into the score: a first category we directly included based on findings in our prior chronic distress metabolomics study that utilized a robust discovery-validation workflow^8^, and a second category that underwent subsequent variable selection using a LASSO conditional logistic regression model (Figure 1, Table S1).

Following this strategy resulted in:

(i) 10 out of 11 metabolites (Table S1) validated in Shutta, KH et al. (2021) ^8^ were included into the MDS. An exception was made for hydroxycotinine excluded due to its high correlation with cotinine.

(ii) 89 metabolites (Table S1) identified in step c) were considered as candidates for inclusion into the MDS. To reduce collinearity and create a more parsimonious score, all 89 metabolites in the NHS sample were input simultaneously into a Lasso conditional logistic regression model adjusting for matching factors and with chronic distress case/control status as the outcome. Each metabolite regression coefficient was assigned a p-value based on a permutation test. Metabolites with permutation p-value and FDR p-value < 0.05 were selected for inclusion in the MDS (N=11).

This process resulted in 21 metabolites identified as eligible for inclusion in the MDS; of these, 20 were available in the WHI-OS and 17 available in PREDIMED. Consequently, two versions of the MDS were derived in the NHS, referred to as the 20-MDS and 17-MDS, respectively. We estimated coefficients of the component metabolites in each MDS in minimally adjusted, unpenalized multivariable conditional logistic regression models, with chronic distress case/control status as the outcome, thereby training the MDS against the phenotype measure.

#### Stage 2: Assessing associations of the metabolite-based distress score with incident CHD and CHD risk factors in the WHI-OS

For each participant in the nested case-control study of CHD in the WHI-OS (n=944), the 20-MDS was calculated as:

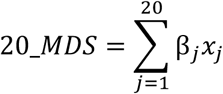

where *β*_*j*_is the coefficient of metabolite *j* estimated in the conditional logistic regression model with chronic distress as the outcome in the NHS, and *x*_*j*_is the log-transformed, standardized metabolite value observed for metabolite *j* in the WHI-OS.

Using logistic regression models, we assessed the association of the 20-MDS assessed at study baseline with odds of incident CHD assessed over 16.7 mean years of follow-up^6^ in three models:

i. a minimal model consisting of the 20-MDS and matching factors (age, race/ethnicity, hysterectomy status and time period of enrollment)
ii. an intermediate model consisting of the 20-MDS, matching factors, and key covariates selected based on prior work examining distress in relation to CHD^1^ as well as to mirror those used in analyses conducted for the parent WHI CHD metabolomics investigation by Paynter, NP et al (2018)^9^. Covariates were measured at baseline and included medications (all yes/no; hypertension treatment, diabetes treatment, aspirin use, statin use), measured systolic blood pressure (SBP), smoking status, and prevalent diabetes status (yes/no).
iii. a full model consisting of all covariates in the intermediate model plus baseline measures of total cholesterol, high-density lipoprotein (HDL) cholesterol. We fit models (ii) and (iii) separately because several lipids were included in the MDS, and we expected the score would be highly correlated with total and HDL cholesterol. Comparing models (ii) and (iii) would allow us to estimate the extent to which the association between MDS and CHD is attenuated by total and HDL cholesterol. To assess whether there is a time-varying association of the 20-MDS measured at baseline with subsequently developing CHD, we performed a stratified analysis, fitting models (i)-(iii) above separately among early-occurring CHD cases (time to CHD < 6 years) and later-occurring CHD cases (time to CHD >= 6 years). Lastly, each metabolite component of the 20-MDS was evaluated individually with odds of incident CHD in logistic regression models with minimal (i) and full risk factor adjustment (iii) as described above. Metabolites satisfying q value < 0.05 in the minimal model and p value < 0.05 in the full model were considered significant associations.

As a secondary analysis, we also fit a separate linear regression model to examine the cross-sectional association of the 20-MDS with the Framingham Risk Score for 10-year CVD risk (outcome)^13^. Three models were used:

- first, a model accounting for matching factors only (age, race, hysterectomy status, time period of enrollment into the WHI-OS);
- second, a model accounting for likely true potential confounders adjusted matching factors along with medications (aspirin use, statin use, anti-diabetic use, anti-hypertensive use); and
- third, a model accounting for biobehavioral factors that could either confound or mediate the associations of interest adjusted for covariates from the second model along with diet ^23^, total recreational physical activity, alcohol intake and BMI.

#### Stage 3: Replication analyses: Assessing association of metabolite-based distress score with total CVD in PREDIMED

For each participant in PREDIMED, the 17-MDS based on 17 component metabolites was calculate similar to the 20-MDS in WHI-OS. Using logistic regression, we evaluated the association of the 17-MDS with risk of incident total CVD ^14^. We performed logistic regression with a similar set of models to those described above for analyses in WHI-OS. These included

i. a minimal model consisting of the 17-MDS as the primary predictor and adjusting for age, sex, and dietary intervention arm,
ii. an intermediate model with additional adjustment for elevated cholesterol, hypertension, diabetes, smoking, and SBP, and
iii. a full model including all covariates in the second model plus total and HDL cholesterol. As PREDIMED includes both male and female participants, we also conducted sex-stratified analyses to evaluate if the relationship between the 17-MDS and CVD is similar across men and women. Following the same strategy used for analyses examining individual metabolites with CVD risk in the WHI-OS, we also tested associations of the individual metabolites of the 17-MDS with incident CVD risk, overall and in sex-stratified analyses. Finally, we investigated the cross-sectional relationship between the 17-MDS and the FRS using a minimally adjusted linear regression of the FRS against the 17-MDS, age, sex, and dietary intervention arm. We further conducted sex-stratified analyses for associations with FRS among women and men, adjusting for age and dietary intervention arm.

## Results

Characteristics of participants in the NHS, WHI-OS and PREDIMED cohorts are shown in Table 1. In PREDIMED, 57% of the sample was women, whereas the NHS and WHI-OS samples are all women. The mean age between cohorts varied between 64-69 years and the proportion non-White was 17% in the WHI-OS, 0% in PREDIMED and 2% in NHS. A few notable differences in participant characteristics across cohorts included higher rates of type 2 diabetes in PREDIMED, lower rate of smoking and higher levels of physical activity and HT use in the NHS (Table 1). The two variations of the MDS derived in the NHS as described in the Methods, namely the 20-MDS and 17-MDS, included 16 metabolites in common (Table 2). N2, N2-dimethylguanosine, serotonin, arachidonate and 3-methylxanthine were included only in the 20-MDS as they were measured in the WHI-OS but not PREDIMED; dimethylguanidino valerate (DMGV) was included only in 17-MDS as this was measured in PREDIMED, but not the WHI-OS.

### Associations of the MDS with incident CHD and CHD risk factors in the WHI-OS

The 944 participants in the WHI-OS included 472 CHD cases with a mean time to CHD of 5.8 years from the time the MDS was assessed. The median (2.5^th^ – 97.5^th^ percentile) of the 20-MDS among CHD cases was 0.08 (−2.3, 3.5) and -0.25 (−2.6, 2.6) among controls (Figure S1). A higher 20-MDS was associated with increased incident CHD risk in the minimal model, with an OR of 1.22 (95% CI: 1.10 – 1.34,) corresponding to a 1 SD increase in the MDS score. In the intermediate model the OR for incident CHD per SD increase in score was 1.14 (95% CI: 1.03 – 1.26). In the full model, the association of 20-MDS with incident CHD was attenuated, with an OR of 1.09 (95% CI: 0.98 – 1.21). See Table 3.

**Table 3:** Metabolite-based distress score derived in the Nurses’ Health Study (NHS) and evaluated in (a) Women’s Health Initiative Observational Study (WHI-OS) for association with CHD; and (b) Prevención con Dieta Mediterránea (PREDIMED) with total CVD. **M1:** In the WHI, adjusted for matching factors including age, race, hysterectomy status and time period of enrollment, assessed at baseline. **M2:** In the WHI, M1 + hypertension treatment, diabetes treatment, aspirin use, statin use, SBP, smoking and diabetes, assessed at baseline. **M3:** In the WHI, M2+ total and HDL cholesterol, assessed at baseline. **M4:** In PREDIMED, adjusted for age, sex and indicator of the three diet intervention arms, assessed at baseline. **M5:** In PREDIMED, M4+ elevated cholesterol, hypertension, diabetes, smoking, SBP, assessed at baseline. **M6:** In PREDIMED, M5 + total cholesterol, HDL cholesterol, assessed at baseline. **M4*:** In PREDIMED, M4 excluding sex. **M6*:** In PREDIMED, M6 excluding sex.

| Outcome | Model | OR (95% CI)<br>(for 1 SD MDS increase) | P value |
| --- | --- | --- | --- |
| **Women's Health Initiative Observational Study** |  |  |  |
| CHD (all events, # cases=472) | M1 | 1.22 (1.10 – 1.34) | 7.03X10<sup>-5</sup> |
|  | M2 | 1.14 (1.03 – 1.26) | 0.01 |
|  | M3 | 1.09 (0.98 – 1.21) | 0.12 |
| CHD<br>(events within 6 years, cases=232) | M1 | 1.24 (1.10 – 1.40) | 6.1x10<sup>-4</sup> |
|  | M2 | 1.12 (0.98 – 1.28) | 0.09 |
|  | M3 | 1.07 (0.93 – 1.22) | 0.35 |
| CHD<br>(beyond 6 years, cases=240) | M1 | 1.20 (1.07 – 1.35) | 1.9x10<sup>-3</sup> |
|  | M2 | 1.14 (1.01 – 1.29) | 0.03 |
|  | M3 | 1.10 (0.97 – 1.25) | 0.13 |
| **PREDIMED** |  |  |  |
| CVD<br>(all, # cases=224) | M4 | 1.24 (1.07 – 1.43) | 0.01 |
|  | M5 | 1.17 (1.00 – 1.36) | 0.05 |
|  | M6 | 1.17 (1.00 – 1.38) | 0.05 |
| CVD<br>(women, # cases=88) | M4* | 1.21 (0.96 – 1.53) | 0.11 |
|  | M6* | 1.12 (0.87 – 1.44) | 0.38 |
| CVD<br>(men, # cases=136) | M4* | 1.25 (1.03 – 1.52) | 0.02 |
|  | M6* | 1.22 (0.98 – 1.51) | 0.08 |

The association of 20-MDS with early-occurring CHD events (within 6 years of blood draw) was similar to that with later events occurring beyond 6 years (Table 3). Of the 20 metabolites comprising the 20-MDS, lower levels of threonine (OR=1.22, 95% CI: 1.06 – 1.42) and tryptophan (OR=1.16, 95% CI: 1.01 – 1.33) were each associated with increased risk of developing CHD risk in the WHI-OS (Figure 2, Figure S2, Table S2) and had consistent directions of associations with chronic distress in the NHS. Four other metabolites were associated with incident CHD risk in the minimal model only – pseudouridine, N2,N2-dimethylguanosine and C16:0 ceramide had positive associations and biliverdin had an inverse association (Figure S2, Table S2).

**Figure 2:**
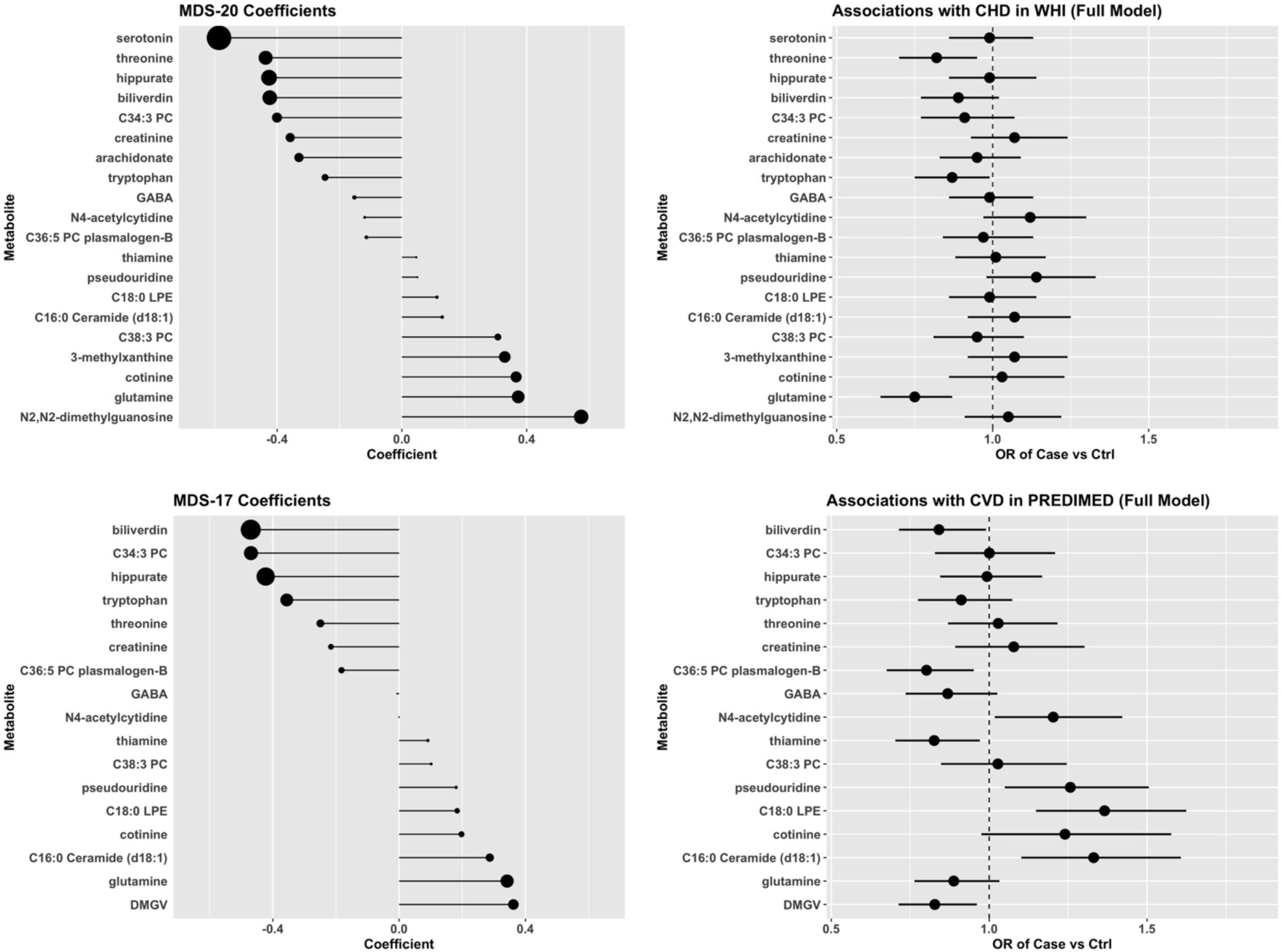
Coefficients s of each in the metabolite-based distress score (20-MDS, 17-MDS) along with individual metabolite associations with incident CHD in the WHI-OS, and with incident total CVD in PREDIMED. <u>NHS</u>: models adjusted for matching factors including baseline assessments of age, race/ethnicity (white/non-white), fasting status (yes/no), menopausal status (yes/no), and date and time of blood draw. <u>PREDIMED</u>: models adjusted for study design factors (age at baseline, sex, and dietary intervention arm), co-morbidities (elevated cholesterol, hypertension, diabetes), total cholesterol, HDL cholesterol, smoking, and SBP, assessed at baseline. <u>WHI-OS</u>: models adjusted for matching factors (age at baseline, race, hysterectomy status at baseline and time period of enrollment into the study), medications (hypertension treatment, diabetes treatment, aspirin, statins), measured systolic blood pressure (SBP), smoking status, and prevalent diabetes status, total and HDL cholesterol, assessed at baseline.

In a cross-sectional analysis, a 1 SD increase in the 20-MDS was associated with a 0.10 (95% CI: 0.06 – 0.12) unit increase in FRS for 10-year CVD risk, after adjusting for age, race/ethnicity, hysterectomy status and time period of enrollment (Table 4). With further adjustment for medication use and biobehavioral factors may link distress to CVD, the association was attenuated but remained statistically significant with a 1 SD increase in 20-MDS associated with a 0.03 (95% CI: 0.01 – 0.06, p =0.01) unit increase in FRS.

**Table 4:** Association between MDS and Framingham risk score (FRS) for 10-year CVD risk in the WHI-OS and PREDIMED. FRS is calculated based on age, total cholesterol, HDL cholesterol, systolic blood pressure (treated or untreated), smoking status and diabetes in a sex-specific algorithm described in D’Agostino et al. (2008)^12^.

| Cohort | Strata | Model | Mean change in FRS per 1<br>SD change in MDS<sup>3</sup><br>(95% CI) | P value |
| --- | --- | --- | --- | --- |
| WHI-OS<sup>4</sup> | All | M1 | 0.09 (0.06 – 0.12) | 6.8x10<sup>-10</sup> |
|  | All | M2 | 0.05 (0.02 – 0.07) | 0.0001 |
|  | All | M3 | 0.04 (0.02 – 0.06) | 0.002 |
| PREDIMED<sup>5</sup> | All | M4 | 0.10 (0.06 – 0.13) | 5.5x10<sup>-9</sup> |
|  | Women | M4* | 0.08 (0.04 – 0.13) | 5.0x10<sup>-4</sup> |
|  | Men | M4* | 0.11 (0.07 – 0.15) | 1.2x10<sup>-6</sup> |

<sup>3</sup>WHI-OS: 20-MDS; PREDIMED: 17-MDS
<sup>4</sup>In the WHI-OS, the models fit were:
M1: adjusting for matching factors including age at baseline, race, hysterectomy status and time period of enrollment.
M2: M1+medications (statin, aspirin, anti-diabetic, anti-hypertensive) assessed at baseline.
M3: M2+bio-behavioral factors (diet assessed by the Healthy Eating Index (HEI))<sup>27</sup> + total recreational physical activity + alcohol intake + BMI) assessed at baseline.
<sup>5</sup> In PREDIMED, the model M4 adjusts for age at baseline, sex and dietary intervention arm. The model M4 adjusts for age and baseline and dietary intervention arm.

### Replication: Associations of the MDS with incident CHD and CHD risk factors in PREDIMED

Of the 980 participants in PREDIMED, 224 experienced incident CVD (mean time to CVD of 3.1 years). Each one SD increase in 17-MDS was associated with increased incident CVD risk with an OR of 1.17 (95% CI: 1.00 – 1.38), after fully adjusting for all risk factors (Table 3). The association of 17-MDS with incident CVD risk was similar in men and women (Table 3). Of the 17 separate metabolites comprising the 17-MDS, 4 were associated with incident CVD risk in fully adjusted models and with consistent directions of associations with chronic distress in the NHS. Biliverdin and C36:5 PC plasmalogen had inverse associations with CVD risk. C16:0 ceramide and C18:0 LPE each had positive associations with CVD risk (Figure 2, Figure S2, Table S3). An increase of 1 SD in the 17-MDS was also associated with a 0.1 (95% CI: 0.06 – 0.13, p < 10^−8^) unit increase in FRS for 10-year CVD risk at baseline after adjusting for age, sex and intervention arm. The association of 17-MDS with FRS was also comparable in men and women, with mean increase in risk score of 0.08 (95% CI: 0.04 – 0.13, p<10^−3^) in women and 0.11 (95% CI: 0.07 – 0.15, p <10^−5^) in men (Table 4).

## Discussion

In the current study, we derived metabolomic profiles of chronic distress that included 20 (20-MDS) and 17 (17-MDS) metabolites, respectively. In a prospective evaluation of women in the WHI-OS, we then found a higher 20-MDS score was associated with increased incident CHD risk after adjusting for matching factors, medication use, co-morbidities and biobehavioral factors. However, this association was attenuated and no longer reached statistical significance after including total and HDL cholesterol in the model. Also, in the WH, a higher 20-MDS was significantly associated with increased 10-year risk of CVD. We replicated these findings in an independent and prospective evaluation that included both men and women; the 17-MDS was significantly associated with incident total CVD risk in 980 men and women participating in PREDIMED, in a model adjusting for all relevant covariates. Similarly, using PREDIMED data we replicated the finding that the 17-MDS was cross-sectionally associated with increased 10-year risk of CVD.

Our findings shed light on the metabolomic underpinnings of previously well documented associations of common mental disorders including depression, anxiety and related conditions with increased incident cardiovascular disease risk ^1^. To gain further insight into these findings, we focus our discussion on the individual components of the MDS that were associated with incident CHD or CVD risk in the WHI-OS or PREDIMED after full adjustment for potential confounders – these included amino acids threonine and tryptophan, biliverdin and a glycerophospholipid (C36:5 PC plasmalogen) that had inverse associations with distress and with CHD/CVD and two lipids (ceramide (C16:0), C18:0 LPE) that had positive associations with CHD/CVD.

Threonine is an essential L-alpha amino acid and active in several biochemical pathways. Lower threonine levels were associated with chronic distress in the NHS and with increased CHD risk in the WHI-OS. There are limited prior reports of associations of threonine with distress. A cross-sectional study on 64 participants reported higher threonine levels among participants with depression and anxiety relative to healthy controls^24,25^, but a prospective study in 841 Japanese participants failed to find an association between initial threonine levels and subsequent incidence of depressive symptoms^26^. In other studies, lower plasma threonine levels have been linked with higher cardiometabolic risk. For example, in a cross-sectional study of 475 individuals, plasma threonine levels were inversely associated with an atherogenic lipid profile ^27^.

In the current study, tryptophan was inversely associated with both chronic distress in NHS and with incident CHD in the WHI-OS, respectively. Tryptophan, an indolyl carboxylic acid and an essential amino acid, is well known as the biochemical precursor to the neurologically active compounds serotonin and melatonin. As tryptophan is the precursor to dietary serotonin (5-hydroxytryptamine or 5-HT), experimental studies of the role of 5-HT in cognitive function and psychiatric disorders show that acute tryptophan depletion results in transient depressive relapse in patients experiencing remission of symptoms after antidepressant therapy ^28 29^. Prior cross-sectional studies also confirm our findings, including a small Chinese study of major depressive disorder (MDD) ^30^ and a larger German study of type D personality characterized by negative affectivity and social isolation ^31^. Previous reports in the literature have also confirmed the role of altered tryptophan levels with CVD risk. Proinflammatory cytokine interferon-*γ*that plays a major role in atherosclerosis activates the enzyme indoleamine 2,3-dioxygenase (IDO), which in turn depletes serum levels of tryptophan^32^. In a previous publication using data from PREDIMED, increases in plasma tryptophan levels were associated with decreased incident CVD risk ^33^.

Biliverdin, a metabolite in the class of bilirubins, was inversely associated with chronic distress and with incident CHD in the WHI-OS and incident CVD in PREDIMED. These observations are consistent with the limited prior data on biliverdin. In a small cross-sectional Polish study, the enzyme heme oxygenase 1, which catalyzes degradation of heme to biliverdin, iron and carbon monoxide ^34-37^, was significantly lower among those depressed and negatively correlated with severity of depressive symptoms^34^.. Biliverdin is itself an antioxidant; our observation of an inverse association with distress is concordant with prior work linking elevated oxidative stress with psychosocial distress^38^. Consistent with our findings, previous studies have found inverse associations of bilirubin (which is derived from biliverdin) with cardiometabolic disorders^39-41^.

Plasma levels of C36:5 PC plasmalogen were inversely associated with both chronic distress in NHS? and with incident CVD risk in PREDIMED. Plasmalogens are glycerophospholipids, are found as components of cell membrane structures and some have anti-oxidant properties ^42^. Prior work has found higher stress associated with decreases in plasmalogens ^43^. For example, a randomized, controlled trial of a lifestyle intervention in a population with high levels of perceived stress reported increases in other PC plasmalogens (18:0/22:6, 18:0/20:4) in the intervention arm and inverse correlations with a heart rate variability based stress index^44^. Although data are limited, inverse associations of PC plasmalogens (including C35:5) with CVD risk have also been reported in both cross-sectional ^45^ and longitudinal studies ^46^.

Higher plasma levels of a ceramide (C16:0) were associated with chronic distress in NHS and with increased CHD/CVD risk in the WHI-OS and PREDIMED. Ceramides are sphingolipids that include circulating and membrane bound lipids composed of a fatty acid linked with a long sphingosine carbon backbone; they are products of lipid metabolism that accumulate in individuals with dyslipidemia. Prior work has found cross-sectional associations of ceramides with chronic distress. For example, a small of 46 participants recruited from a U.S. memory clinic reported significantly elevated ceramides levels (including 16:0 ceramide) among those with major depression, irrespective of dementia status^47^. Similarly, the role of ceramides and other sphingolipids in cardiometabolic disorders have been demonstrated ^48 48-53^. Such work has indicated that accumulation of circulating lipids including ceramides in blood vessels and the heart results in vascular/cardiac dysfunction and contributes to cardiometabolic disease ^48,54^.

Another lipid, C18:0 LPE, a lysophospatidyl ethanolamine, was significantly elevated in the distressed cases relative to controls in our study. A positive association with incident CVD risk was also observed in PREDIMED. Lipids including LPEs are essential components of the cell membrane, with key roles in signal transduction and synaptic activity. Previous studies have linked alterations in lipid metabolism to depression. For example, a cross-sectional Chinese study of 119 participants reported elevated levels of plasma 18:2 LPE in those with major depressive disorder (MDD) relative to healthy controls^30^. Associations of plasma lipid classes with incident CVD risk have been previously reported in multiple longitudinal cohorts ^55-57 58^.

To our knowledge, this is one of the first studies to directly derive a metabolite-based distress score and examine its association with future risk of CVD-related endpoints. Our metabolite-based distress score was developed in cohorts of women only, but appear to be relevant in men as our findings linking the distress-based metabolite score with CVD were consistent across men and women. Relatedly, the primary association of our MDS with CVD and CVD risk factors replicated across two completely different cohorts. Findings from the current study provide important insights into key mechanisms that may underlie long-observed associations between various forms of psychological distress and endpoints related to CVD. We identified metabolite alterations that reliably occur in conjunction with psychological distress and also found evidence that these metabolite alterations in combination are associated with subsequent risk of developing cardiovascular disease and with a poorer profile across a range of known CVD risk factors. Future work will be needed to assess the direction of associations between distress and these metabolite alterations, to determine more conclusively if distress truly precedes and contributes to substantial metabolomic dysregulation that in turn leads to greater risk of CVD. If these findings hold, they suggest such dysregulation may also be likely to explain observed associations of psychological distress with increased risk of stroke or diabetes more specifically ^2,3,59^. Important additional questions to explore with regard to these findings include what duration of distress is needed to drive metabolic dysregulation and how early in the life course are such alterations evident. It will also be important to evaluate if these associations are similarly evident in more diverse populations and if there are additional metabolites (as yet unidentified) that might contribute meaningfully to our metabolite-based distress score.

Our study has some limitations. First, the derivation of MDS was carried out with cross-sectional data; second, the three cohorts (NHS, WHI-OS, PREDIMED) are sampled from distinct populations, thereby introducing systemic variability into our analyses; third, the MDS was derived in the NHS that included post-menopausal women, with limited racial/ethnic diversity. However, our study has a number of unique strengths, including the use of multiple, richly characterized cohorts as well as the wide range of small molecule metabolites measured on each sample using the same metabolomics platforms. Our evaluation of the association between the metabolite-based distress score and subsequent risk of incident CHD or CVD was conducted in prospective investigations in two independent, diverse cohorts of similar ages. Moreover, sex-stratified analyses in PREDIMED demonstrated that the MDS is similarly associated with CVD in both men and women.

In summary, our study identified a metabolomic score of chronic distress that was significantly associated with future risk of CVD in two independent cohorts, including replication in men and women. These metabolites reflect a diverse set of metabolic processes including pathways serving as precursors to neurologically active compounds, anti-inflammatory and inflammatory processes including adipose tissue inflammation and vascular dysfunction, and those involving accumulation of neurotoxic compounds and nicotine metabolism. A clearer understanding of key molecular alterations that may result from chronic distress may provide stronger evidence that distress indeed causally contributes to greater risk of developing CVD and also give greater insight into mechanisms by which these effects occur. Further exploration of these mechanisms might provide critical insight regarding whether effects of chronic distress on CVD are reversible or can be disrupted in meaningful ways to break the chain of risk and promote cardiovascular health over the life course.

## Data Availability

The WHI metabolomics data are available for request through dbGAP (dbGaP Study Accession: phs001334.v1.p3). Other data included in this study are available upon reasonable request to the authors.

## Sources of Funding

The NHS is supported by the National Institutes of Health (UM1CA186107, R01CA49449). Research reported in this publication is supported by the National Institutes of Health under award number R01AG05160001A1. Metabolomic analysis in the WHI was funded by the National Heart, Lung, and Blood Institute, National Institutes of Health, U.S. Department of Health and Human Services through contract HHSN268201300008C. The WHI program is funded by the National Heart, Lung, and Blood Institute, National Institutes of Health, U.S. Department of Health and Human Services through 75N92021D00001, 75N92021D00002, 75N92021D00003, 75N92021D00004, 75N92021D00005. A list of WHI investigators is available online at https://www-whi-org.s3.us-west-2.amazonaws.com/wp-content/uploads/WHI-Investigator-Short-List.pdf.

## Disclosures

None

## Notes

### Competing Interest Statement

The authors have declared no competing interest.

### Author Declarations

IRB of the University of Massachusetts Amherst gave ethical approval for this work (IRB protocol number 2017-4095, titled "Development and Application of Metabolomic Profile of Chronic Distress to Cardiometabolic Risk"

