## Supplemental Material for "Metabolomic profiles of chronic distress predict future cardiovascular disease risk"

### Study Populations

NHS: NHS participants had their blood drawn in sodium heparin tubes and shipped with an ice pack via overnight courier to the laboratory, where it was processed into plasma, red blood cells and white blood cells[16]. Blood samples were divided into small aliquots and were stored at -130°C or colder in the vapor phase of liquid nitrogen freezers.

WHI-OS: Baseline fasting blood samples were collected and processed at local centers into separate aliquots containing serum, plasma and buffy coat, which were frozen and shipped to a central repository for long-term storage[17]. Samples were divided into small aliquots and were stored at -70°C in mechanical freezers since collection and until analysis

PREDIMED: Plasma samples were collected using EDTA and processed immediately[17]. Samples were divided into small aliquots and were stored at -70°C in mechanical freezers since collection and until analysis.

### Distress Definition and Case/Control Selection in the NHS

The process described below was used to select cases and controls in the NHS validation dataset. Here, "concurrent" refers to an assessment that was at or near the time of the blood draw, and "prior" refers to an assessment that was taken well before the blood draw.

Levels of depression were ascertained using the Mental Health Inventory (MHI-5) [1,2], assessed in 1992, 1996, and 2000, and CES-D-10[3], assessed in 2004, as well as reports of physician-diagnosed depression, assessed in 2000 and 2004, and use of antidepressants, assessed in 1996, 2000, and 2004. Levels of anxiety were ascertained using the phobic anxiety scale of the Crown Crisp Experimental Index (CCI) in 1988 and 2004, which discriminates individuals with diagnosable anxiety disorders from healthy individuals and correlates reasonably with other measures of anxiety [4] and the validated State-Trait Anxiety Inventory (STAI)[5].

Prior depressive episodes were defined as occurring at each of those times if participants were positive on 1 or more of the following: MHI-5 score<=52 or CES-D-10 score >= 10,[2] antidepressant use, or physician-diagnosed depression at the time of the relevant questionnaire. Concurrent depression was derived from the 2004 questionnaire and defined as occurring if participants were positive on 1 or more of the following: CES-D-10 score >= 10, antidepressant use, or physician-diagnosed depression. We defined individuals with *persistent depression* as those with concurrent depression in 2004 and who also reported experiencing depressive episodes at least twice across the 3 time points of 1992, 1996 and 2000. We defined individuals with *episodic depression* as those with concurrent depression in 2004 and who reported experiencing a depressive episode in at most one prior assessment. Anxiety was assessed on the 1988 questionnaire using the CCI and on the 2004 questionnaire using the STAI. Following earlier work in this cohort[6], prior anxiety was defined as reporting ≥ 3 symptoms (out of 8 symptoms total) on the CCI. Concurrent anxiety was defined by an STAI score ≥ 22[7] on the 2004 questionnaire. Women with both concurrent and prior anxiety at both times were considered to have *persistent anxiety*.

To capture women with chronic and more severe distress manifested as either depression and/or anxiety, we carried out the following sampling strategy to obtain 280 cases with chronically high distress and an equal number of matched controls with low levels of distress at every time point where measurements were available. Cases were obtained by selecting (1) all women with both persistent depression and persistent anxiety (n=176); (2) an additional 104 women from among (a) 117 women with persistent depression and current anxiety and (b) 180 women with persistent anxiety and episodic depression, oversampling those with high current depression or current anxiety symptoms. The control pool included those with no current or past depression or anxiety according to the same above definitions at any of the 4 timepoint (n=9,221 of 13,194 eligible participants).

**Details on outcome and covariate assessment**

**WHI-OS:**

Blood pressure was measured by certified staff in the right arm with a mercury sphygmomanometer after the participant was seated and had rested for 5 minutes[22,23]. The average of 2 blood pressure readings obtained at least 30 seconds apart, was used for analysis [22,23].

Medication use, smoking, diet and physical activity: Information on medication use was collected by study staff at the baseline clinic visit by inventory of medication and supplement pills[24]. Information on smoking status and co-morbidities were self-reported on the lifestyle and medical history questionnaires collected at study entry[24]. Diet was assessed with an Food Frequency Questionnaire (FFQ)[25], from which an overall Healthy Eating Index (HEI-2005) score[26] was derived along with measurements of alcohol (drinks/day) intake. Activity in MET-hours per week was calculated based on WHI brief recreational physical activity inventory (https://www.whi.org/dataset/31).

**Metabolomics assay**

In NHS, participants had their blood drawn in sodium heparin tubes at a nearby clinic and shipped with an ice pack via overnight courier to the laboratory, where it was processed into plasma, red blood cells and white blood cells (Huang et al. 2019). In the WHI-OS, baseline fasting blood samples were collected and processed at local centers into separate aliquots containing serum, plasma and buffy coat, which were frozen and shipped to a central repository for long-term storage(Anderson et al. 2003). In PREDIMED, plasma samples were collected using EDTA and processed immediately[17]. In all three cohorts, blood samples were divided into small aliquots and were stored at -130$^{o}$C or colder in the vapor phase of liquid nitrogen freezers (NHS) or at -70°C in mechanical freezers (WHI, PREDIMED) since collection.

Internal standard peak areas were monitored for quality control and to ensure system performance throughout analyses. Pooled plasma reference samples were also inserted every twenty samples as an additional quality control. A C8-positive platform was used to measure polar and non-polar plasma lipids, identified on the basis of total acyl carbon content and degrees of saturation, including triacylglycerols, diacylglycerols, cholesterol esters, sphingomyelins, phosphatidylcholines, and lysophospholipids. A HILIC-positive platform was used to measure water soluble metabolites, including amino acids, amino acid derivatives, amines and other metabolites. Negative ion mode analyses were conducted to measure free fatty acids and bile acids (C18-neg) using an LC-MS system. Targeted data were processed using MultiQuant software (AB SCIEX; Framingham, MA) or TraceFinder (Thermo Fisher Scientific; Waltham, MA) for automated LC-MS peak integration.

Metabolomic measurements were made using three complimentary LC-MS methods. For each method, pooled plasma reference samples were included every 20 samples and results were standardized using the ratio of the value of the sample to the value of the nearest pooled reference multiplied by the median of all reference values for the metabolite.

HILIC analyses of water-soluble metabolites in the positive ionization mode (HILIC-pos) were conducted using an LC-MS system comprised of a Shimadzu Nexera X2 U-HPLC (Shimadzu Corp.; Marlborough, MA) coupled to a Q Exactive hybrid quadrupole orbitrap mass spectrometer (Thermo Fisher Scientific; Waltham, MA). Plasma samples (10 $\mu$L) were prepared via protein precipitation with the addition of nine volumes of 74.9:24.9:0.2 v/v/v acetonitrile/methanol/formic acid containing stable isotope-labeled internal standards (valine-d8, Sigma-Aldrich; St. Louis, MO; and phenylalanine-d8, Cambridge Isotope Laboratories;

Andover, MA). The samples were centrifuged (10 min, 9,000 x g, 4°C), and the supernatants were injected directly onto a 150 x 2 mm, 3 μm Atlantis HILIC column (Waters; Milford, MA). The column was eluted isocratically at a flow rate of 250 μL/min with 5% mobile phase A (10 mM ammonium formate and 0.1% formic acid in water) for 0.5 minute followed by a linear gradient to 40% mobile phase B (acetonitrile with 0.1% formic acid) over 10 minutes. MS analyses were carried out using electrospray ionization in the positive ion mode using full scan analysis over 70-800 m/z at 70,000 resolution and 3 Hz data acquisition rate. Other MS settings were: sheath gas 40, sweep gas 2, spray voltage 3.5 kV, capillary temperature 350°C, S-lens RF 40, heater temperature 300°C, microscans 1, automatic gain control target 1e6, and maximum ion time 250 ms.

Positive ion mode analyses of polar and non-polar plasma lipids (C8-pos) were conducted using an LC-MS system comprised of a Shimadzu Nexera X2 U-HPLC (Shimadzu Corp.; Marlborough, MA) coupled to a Exactive Plus orbitrap mass spectrometer (Thermo Fisher Scientific; Waltham, MA). Plasma samples (10 μL) were extracted for lipid analyses using 190 μL of isopropanol containing 1,2-didodecanoyl-sn-glycero-3-phosphocholine (Avanti Polar Lipids; Alabaster, AL). After centrifugation, supernatants were injected directly onto a 100 x 2.1 mm, 1.7 μm ACQUITY BEH C8 column (Waters; Milford, MA). The column was eluted isocratically with 80% mobile phase A (95:5:0.1 vol/vol/vol 10mM ammonium acetate/methanol/formic acid) for 1 minute followed by a linear gradient to 80% mobile-phase B (99.9:0.1 vol/vol methanol/formic acid) over 2 minutes, a linear gradient to 100% mobile phase B over 7 minutes, then 3 minutes at 100% mobile-phase B. MS analyses were carried out using electrospray ionization in the positive ion mode using full scan analysis over 200–1000 m/z at 70,000 resolution and 3 Hz data acquisition rate. Other MS settings were: sheath gas 50, in source CID 5 eV, sweep gas 5, spray voltage 3 kV, capillary temperature 300°C, S-lens RF 60, heater temperature 300°C, microscans 1, automatic gain control target 1e6, and maximum ion time 100 ms. Lipid identities were determined based on comparison to reference plasma extracts and were denoted by total number of carbons in the lipid acyl chain(s) and total number of double bonds in the lipid acyl chain(s).

Negative ion mode analyses of free fatty acids and bile acids (C18-neg) were conducted using an LC-MS system comprised of a Shimadzu Nexera X2 U-HPLC (Shimadzu Corp.; Marlborough, MA) coupled to a Q Exactive hybrid quadrupole orbitrap mass spectrometer (Thermo Fisher Scientific; Waltham, MA). Plasma samples (30 $\mu$L) were extracted using 90 uL of methanol containing PGE2-d4 (Cayman Chemical Co.; Ann Arbor, MI) and centrifuged (10 min, 9,000 x g, 4°C). The samples were injected onto a 150 x 2 mm ACQUITY T3 column (Waters; Milford, MA). The column was eluted isocratically at a flow rate of 400 $\mu$L/min with 60% mobile phase A (0.1% formic acid in water) for 4 minutes followed by a linear gradient to 100% mobile phase B (acetonitrile with 0.1% formic acid) over 8 minutes. MS analyses were carried out in the negative ion mode using electrospray ionization, full scan MS acquisition over 200-550 m/z, and a resolution setting of 70,000. Metabolite identities were confirmed using authentic reference standards. Other MS settings were: sheath gas 45, sweep gas 5, spray voltage -3.5 kV, capillary temperature 320°C, S-lens RF 60, heater temperature 300°C, microscans 1, automatic gain control target 1e6, and maximum ion time 250 ms.

Raw data from Q Exactive/Exactive Plus instruments were processed using TraceFinder software (Thermo Fisher Scientific; Waltham, MA) and Progenesis QI (Nonlinear Dynamics; Newcastle upon Tyne, UK) while MultiQuant (SCIEX; Framingham, MA) was used to process 5500 QTRAP data. For each method, metabolite identities were confirmed using authentic reference standards or reference samples. CVs were calculated using pooled plasma samples from a subset of the study participants.

**Supplemental Tables**

**Supplemental Table S1**: Metabolites considered for inclusion in the metabolite-based distress scores, 20-MDS and 17-MDS.

File: TableS1.xlsx

**Supplemental Table S2**: Metabolite components of the 20-MDS and associations with incident CHD risk in the WHI.

File: TableS2.xlsx

**Supplemental Table S3**: Metabolite components of the 17-MDS and associations with incident CVD risk in PREDIMED.

File: TableS3.xlsx

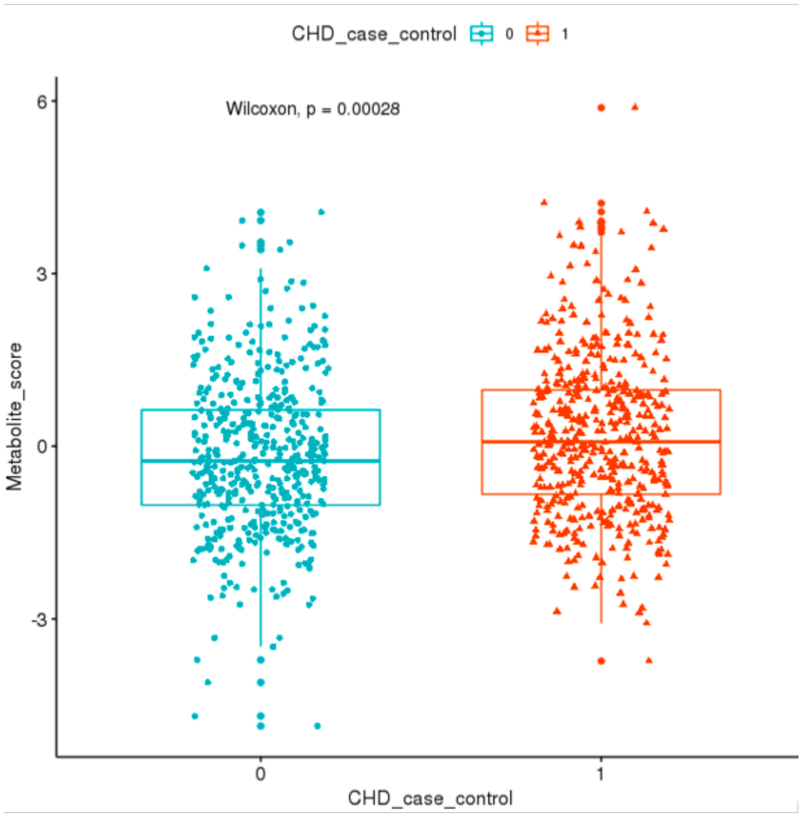


**Figure S1:** Distribution of the 20 metabolite-based distress score (20-MDS) significantly differs between women with CHD and healthy controls in the WHI-OS.

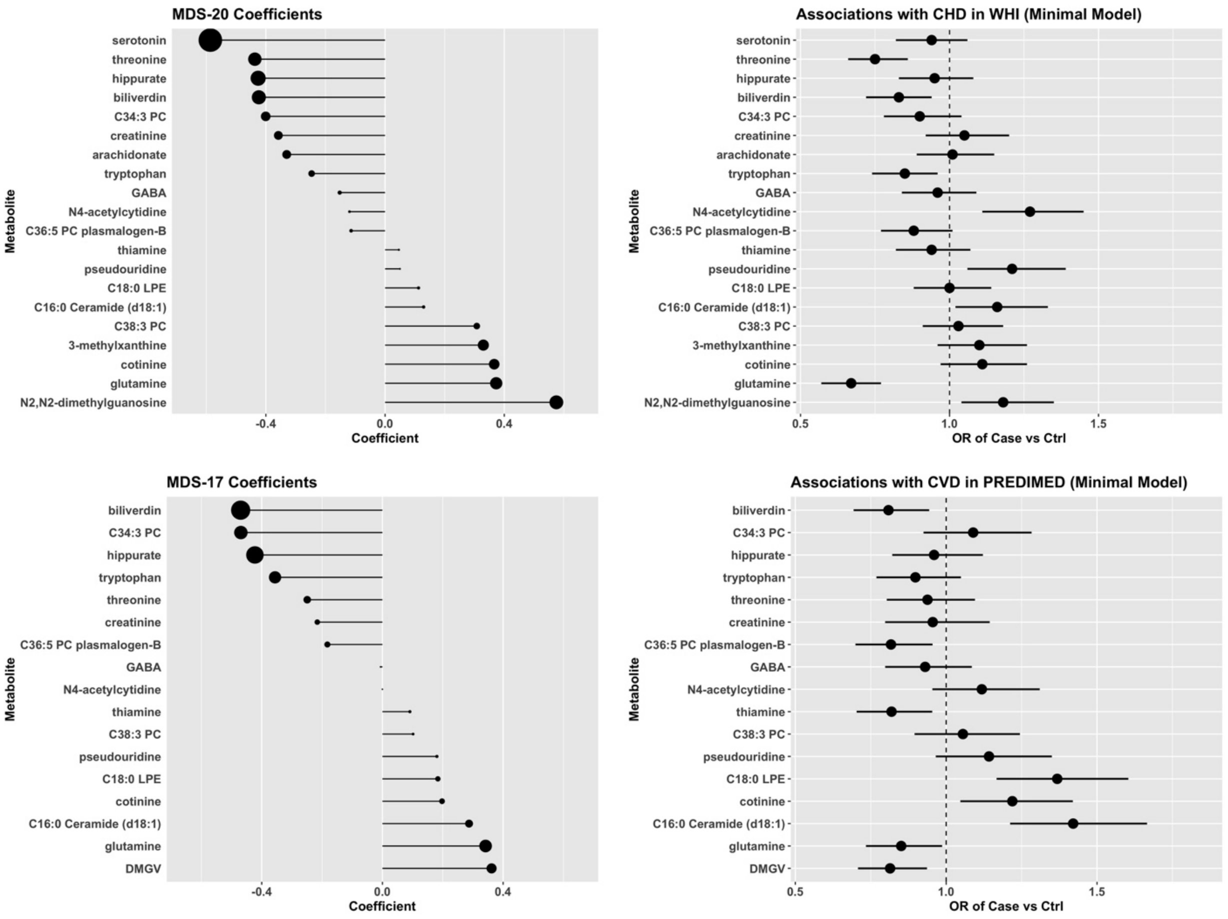


**Figure S2:** Coefficients of individual metabolites in the metabolite-based distress score (20-MDS, 17-MDS) compared to their associations with incident CHD in the WHI and with incident total CVD in PREDIMED. Odds ratios (OR) and associated 95% confidence intervals corresponding to 1 SD increase in metabolite levels are shown.

<u>NHS</u>: metabolite weights in MDS were derived in a conditional logistic regression model that included all metabolite components while adjusting for matching factors including baseline assessments of age, race/ethnicity (non-Hispanic White vs other), fasting status (yes/no), menopausal status (yes/no), and date and time of blood draw.

<u>PREDIMED</u>: models adjusted for study design factors (age at baseline, sex, and dietary intervention arm), assessed at baseline.

<u>WHI</u>: models adjusted for matching factors (age at baseline, race, hysterectomy status at baseline and time period of enrollment into the WHI), assessed at baseline.
